# Brain-gut axis imaging, motion correction with [^11^C]-carfentanil total-body PET

**DOI:** 10.64898/2026.06.17.26355893

**Authors:** Elizabeth J Li, Sophie Lammers, Chia-Ju Hsieh, Joshua Pascale, Joseph Chang, Erin Schubert, Hsiaoju Lee, Robert H. Mach, Joel S. Karp, Corinde Wiers, Henry R. Kranzler, Jacob Dubroff

## Abstract

**Background:** Mu-opioid receptors (MORs) are expressed throughout the body including in the brain and gastrointestinal (GI) tract. Total-body PET imaging of the brain and GI tract offers a promising approach for cross-sectional *in vivo* evaluation of the MOR brain-GI axis. However, intestinal motility and bladder filling introduce motion throughout the GI tract over the scan window. Here we establish analysis methodology to account for motion for dynamic imaging of the brain-GI axis, to further characterize peripheral MORs throughout the body and provide a framework for semi-automatic total-body PET modeling.

**Methods:** 4 subjects underwent 90-min dynamic [^11^C]-carfentanil (cfn) total-body PET acquisitions at baseline, after intravenous naloxone (central antagonist) administration, and after orally administered loperamide (peripheral agonist and P-glycoprotein substrate). Thalamic MOR availability was measured using the Logan reference tissue model. Using CT-based segmentation, the GI tract was subdivided into anatomical segments, in addition to other peripheral organs (e.g., liver, psoas muscle). Frame-by-frame semi-automatic motion correction was performed with three distinct reference frames (11-14 min post-injection, p.i., 35-40 min p.i., and 85-90 min p.i.). The performance of these three were compared to manual correction. Compartment modeling and Logan graphical analysis were performed to estimate relevant kinetic parameters (K1, VT, VTLogan).

**Results:** Across the 4 subjects and regions, kinetic parameter estimates were highly correlated (r>0.7) for K1, VT and VT Logan when comparing semi-automatic (reference frame at 35-40 min p.i.) and manual correction. With semi-automatic motion correction, graphical-based estimation of VTLogan in the gastrointestinal tract was significantly decreased with loperamide relative to baseline (p<0.05). As expected, naloxone decreased brain thalamic MOR availability but loperamide did not.

**Conclusions:** With semi-automatic motion correction and [^11^C]-cfn total-body PET, pharmacologic perturbations of MOR brain-GI axis can be quantitatively characterized, reducing the burden of image analysis for these studies.

## Background

Brain mu-opioid receptors (MORs) play a critical role in the neurocircuitry of reward, motivation, and nociception. Their dysfunction has been implicated in addictive disorders, including opioid use disorder (OUD) and alcohol use disorder. However, MORs are found in abundance throughout the body, including in the lumen of the intestine, where their activation leads to reduced gastrointestinal motility and blood flow, and in the case of chronic opioid use, constipation. There is growing interest in the relationship of the brain-gastrointestinal (GI) axis in OUD and other neuropsychiatric disorders. There is evidence that the GI microbiome and endothelial cell tight junctions are disrupted by opioid use [1–3]. This leads to the introduction of pathogens into the bloodstream, and ultimately to increased GI and systemic inflammatory signaling and opioid tolerance [3]. Further, depletion of the intestinal microbiome in mice induces changes in drug-seeking behavior [4, 5]. However, most PET imaging of MORs in humans to date has focused on the brain. Our group recently used [^11^C]-carfentanil ([^11^C]-cfn), a potent and selective MOR agonist, with dynamic, total-body PET, to investigate the whole-body biodistribution of MORs [6]. Dynamic total-body PET may be a valuable technique for cross-sectional *in vivo* evaluation of the MOR brain-GI axis. With total-body PET, high-temporal-resolution dynamic images can be obtained simultaneously from the brain, intestines, and central blood pool, with high signal quality past 4-5 half-lives of carbon-11 [6–8]. However, there are significant barriers to implementation. First, because dynamic PET acquisitions occur over the course of 60 min or more, the image frames are subject to respiratory, digestive, and bladder filling motion artifacts. Second, in addition to the presence of circulating radiometabolites in the blood, as an excretory organ, the gastrointestinal tract will contain hepatobiliary waste – including radiometabolites.

Here we address the first barrier, through development of analysis methodology for quantification of the brain-GI axis, for further characterization of peripheral MORs with total-body PET. In this pilot study, we pharmacologically manipulate MOR GI availability in healthy controls with dynamic [^11^C]-cfn total-body PET at baseline, following loperamide pretreatment, and after naloxone pretreatment. Loperamide, a synthetic opioid that inhibits intestinal motility, secretion, absorption and blood flow without impacting the central nervous system (CNS) [9], is a P-glycoprotein substrate and is thus not considered blood-brain barrier penetrant. Naloxone, which is used for rapid reversal of opioid overdose, is an MOR antagonist that crosses the blood-brain barrier. After CT-based segmentation [10], motion correction methods were compared and then the best performing method was tested on the intestine for quantification of the volume of distribution (VT). We compare these results to thalamic MOR VT to establish the differential effects of naloxone and loperamide in the brain and GI tract.

## Methods

### PET/CT imaging

4 healthy volunteers (2 male, 2 female, 32-44 years old, 61.8-79.1 kg) underwent three dynamic PET/CT acquisitions on the PennPET Explorer [11]. Subjects also underwent T1-weighted magnetic resonance imaging for localization of cortical and subcortical structures. For each PET/CT, a low-dose CT was acquired for attenuation correction. Subjects were then injected with a bolus of 192.0 ± 54.6 MBq (range: 112.6 – 310.0 MBq) [^11^C]-cfn immediately followed by a saline flush (30 ml) and scanned for 90 min. Baseline scans occurred first, followed by a scan 10 ± 13 days later. The third [^11^C]-cfn scan, after loperamide administration, occurred 1.48 ± 0.55 years later. Intravenous naloxone (13 μg/kg) was administered 10-15 min prior to the PET study, while oral loperamide (4 mg) was administered 45-75 min prior to the PET scan.

### Image analysis and motion correction

Brain image post-processing and quantification were performed independently from peripheral analyses, following our prior work [6]. Briefly, the dynamic [^11^C]-cfn image frames were cropped to focus on the brain, rigidly corrected for frame-by-frame motion, and spatially aligned to the T1 image via rigid registration. The automatic anatomical labeling atlas [12] was used to extract the thalamus and occipital lobe cortical surface.

For peripheral, non-CNS organs, the low-dose CT images were segmented [10] to identify peripheral organs. Various volumes of interest (VOIs) of the colon and small intestine were further manually identified from the segmentations. Due to the limited thickness of the intestinal wall (1-5 mm [13]), the intestinal VOIs were whole-segment, rather than wall-specific, and thus included the intestinal contents. The ascending and descending colon were selected for preliminary modeling. These regions are in the retroperitoneum and are fixed to the posterior abdominal cavity wall. Thus, they are less likely to exhibit extreme motion, whereas the duodenum, ileum, jejunum, and pancreas were all expected to exhibit motion artifacts due to respiratory motion, bladder filling, and peristalsis. VOIs of the whole liver and within the descending aorta were also defined.

In one subject across all three conditions, two methods of motion correction were assessed: manual and semi-automatic. Manual motion correction required manual affine registration (6 degrees of freedom) of the VOIs to individual organs, followed by manual removal of any portion of the VOI that showed visual overlap with other organs. Semi-automatic motion correction was performed using the FALCON tool [14]. FALCON performs a frame-by-frame non-rigid motion correction to a reference frame. Multiple reference frames were carefully selected. The first reference frame was frame 21 (11-14 min post-injection, p.i.), which was chosen to maximize the activity in the brain and show stable activity in the lungs. Second, frame 27 (35-40 min p.i.) was selected as a timepoint prior to bladder filling in all scans. And third, the final frame of the 90-min dynamic study, frame 42 (85-90 min p.i.) was used, where bladder volume was largest. Manually-corrected VOIs from the frame of reference were then applied to the motion-corrected dynamic frames for TAC extraction.

As noted above, evaluation of the three different reference frames was evaluated in a single subject, across the three scanning conditions. After evaluation of the results (described in the following section), a single motion correction reference frame was selected for a comparison with manual motion correction in all subjects.

### Kinetic modeling, quantitative assessments

Thalamic distribution volume ratio (DVRThal) was estimated using the Logan reference tissue model [15]. The visual cortex (modified from calcarine) was used as the reference tissue. A fixed k2’ of 0.12 min^−1^ was used from the literature [16, 17], and as per our prior work [6]. To assess the impact of naloxone and loperamide on [^11^C]-cfn plasma availability, the area under the curve (AUC) was calculated using the trapezoidal method. AUC was measured for the full scan duration, the first 30 min, and 30 to 90 min p.i.

For all regions except the thalamus, the standard deviations within the VOIs were compared across motion correction methods. Based on visual inspection of the data, motion mismatches resulted in a higher standard deviation within the VOIs. Average SUV and the sum of squared residuals (SSR) relative to manual motion correction were also assessed to gauge motion correction performance. For region j and N frames:

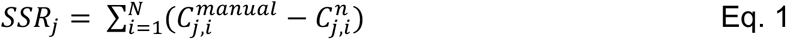

where 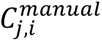 is the average SUV in frame i of the manually-corrected motion dataset, and 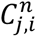 is the semi-automatic motion-corrected dataset, with reference frame n. SSR was then summed across all regions.

To characterize the peripheral [^11^C]-cfn distribution in the GI tract and other organs, one-tissue, two-tissue reversible, and two-tissue irreversible compartment models were implemented using the KMAP software (https://github.com/sharekm). All compartment models included fitting of blood volume fraction and time delay of the bolus entering the tissue space, in addition to the possibility of a dual blood supply for fitting of the liver[18]. Model selection was performed using the Akaike Information Criterion (AIC). After selection of the motion correction reference frame, compartment modeling was performed across all additional subjects and conditions to estimate K1 (mL/min/mL), the volume of distribution (VT, units mL/mL), and the net influx rate of [^11^C]-cfn Ki (mL/min/mL).

In addition, VOI-based and parametric Logan plots [19] were included to assess the feasibility of graphical analysis with implementation of motion correction. For VOI-based analysis, the starting time of the fit (tstar) was allowed to vary from 2 min to 48-min, allowing for a minimum of 9 image frames to be included in the fitting process. Parametric Logan plots of VTLogan were generated with a fixed tstar of 10 min. The volume of distribution (VT) was determined from both compartment modeling and Logan graphical analysis (VTLogan). Based on our prior work [6], the psoas muscle was utilized as a peripheral reference region to estimate the distribution volume ratio (DVR) for both Logan (LoganDVR) and tissue compartment models (TCMDVR), which is equal to the distribution volume in the target tissue divided by the distribution volume of the reference region. To assess the impact of motion correction methods on quantification, modeling results for K1, VT, Ki, TCMDVR and LoganDVR were compared between manual and semi-automatic methods. We report final results based on both the manual and semi-automatic correction methods.

### Statistics

All statistical tests were performed in Matlab (MathWorks, Inc.). Pearson’s correlation coefficient was calculated to evaluate the relationship of parameter estimates across motion correction strategies. The non-parametric Friedman test was used to compare parameters across conditions with *post hoc* analyses using Tukey’s honestly significant difference criterion. A significance threshold of 0.05 was used for all statistical tests.

## Results

Representative brain and body SUV images are shown in **Figure 1**. Visual inspection of the images showed a similar distribution of the tracer across the body, though there were differences in intestinal positioning (arrows). As expected, there was decreased uptake in the brain with naloxone blocking relative to baseline. No visual differences in the brain between baseline and studies with loperamide were observed. Additionally, there were no significant differences in plasma AUC across conditions or time frames.

**Figure 1:**
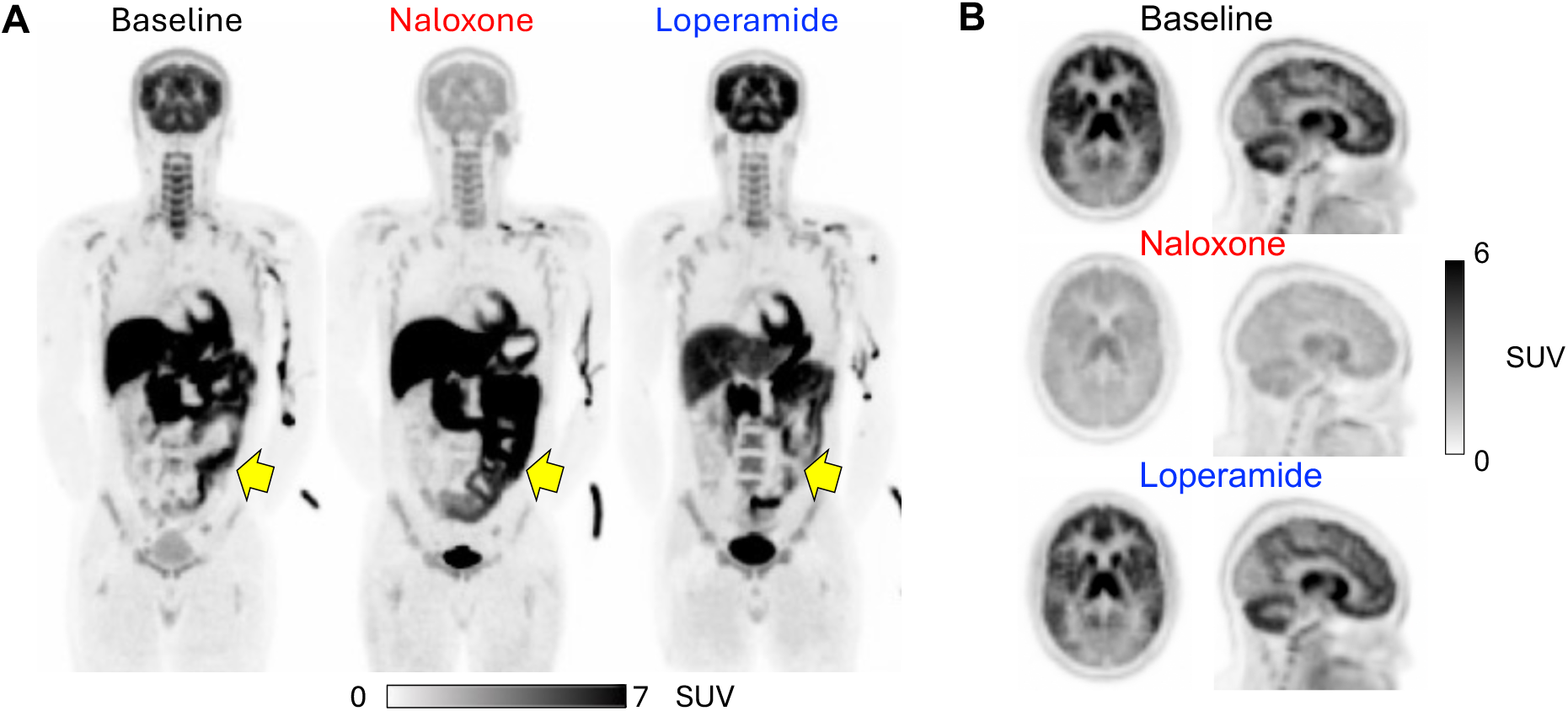
Representative cross-sectional images at baseline, with naloxone, and with loperamide. **(A)**: Coronal images from frame 21 (11-14 min p.i.). Arrows show differences in both intestinal uptake and positioning, motivating the motion correction assessments in this work. **(B)**: Axial and sagittal SUV images of another subject across conditions. SUV was visually lower with naloxone blocking compared to baseline or with loperamide.

### Thalamic occupancy

In the brain, there was a significant difference across groups in thalamic DVR between conditions (p<0.05). More specifically, thalamic DVR was 2.32 ± 0.12 at baseline, 1.60 ± 0.14 with naloxone, and 2.52 ± 0.27 with loperamide. A *post hoc* Tukey test showed that the thalamic DVR was significantly decreased with naloxone blocking relative to loperamide, though the *post hoc* difference between baseline and naloxone was not significant in this small dataset.

### Motion correction

Baseline, naloxone and loperamide scans from a single subject were used to select a reference frame. By visual inspection of the time activity curves and regional standard deviations, manual motion correction outperformed semi-automatic, frame-by-frame motion correction, which was highly dependent on the reference frame selected. **Figure 2** shows retroperitoneal VOIs (ascending and descending colon), duodenum, stomach, and sigmoid as examples. We found that semi-automatic methods performed similarly for retroperitoneal regions. However, semi-automatic methods underestimated the early SUV (<20 min post injection) relative to manual correction, which resulted in the underestimation of peak activity in some tissues (e.g., descending colon, duodenum, **Figure 2**). This underestimation decreased as the reference frame number (and thus time p.i.) increased. VOI standard deviation (shaded region of **Figure 2**) varied with reference frame and organ. Duodenum standard deviation was narrowest with manual correction, particularly in early frames. Manual correction of the stomach resulted in similar results to semi-automatic motion correction when frame 27 or frame 42 was used as the reference. Frame 21 showed a drop in SUV and standard deviation relative to other motion correction methods, while sigmoid standard deviation increased over time with reference frame 21, likely due to bladder filling.

**Figure 2:**
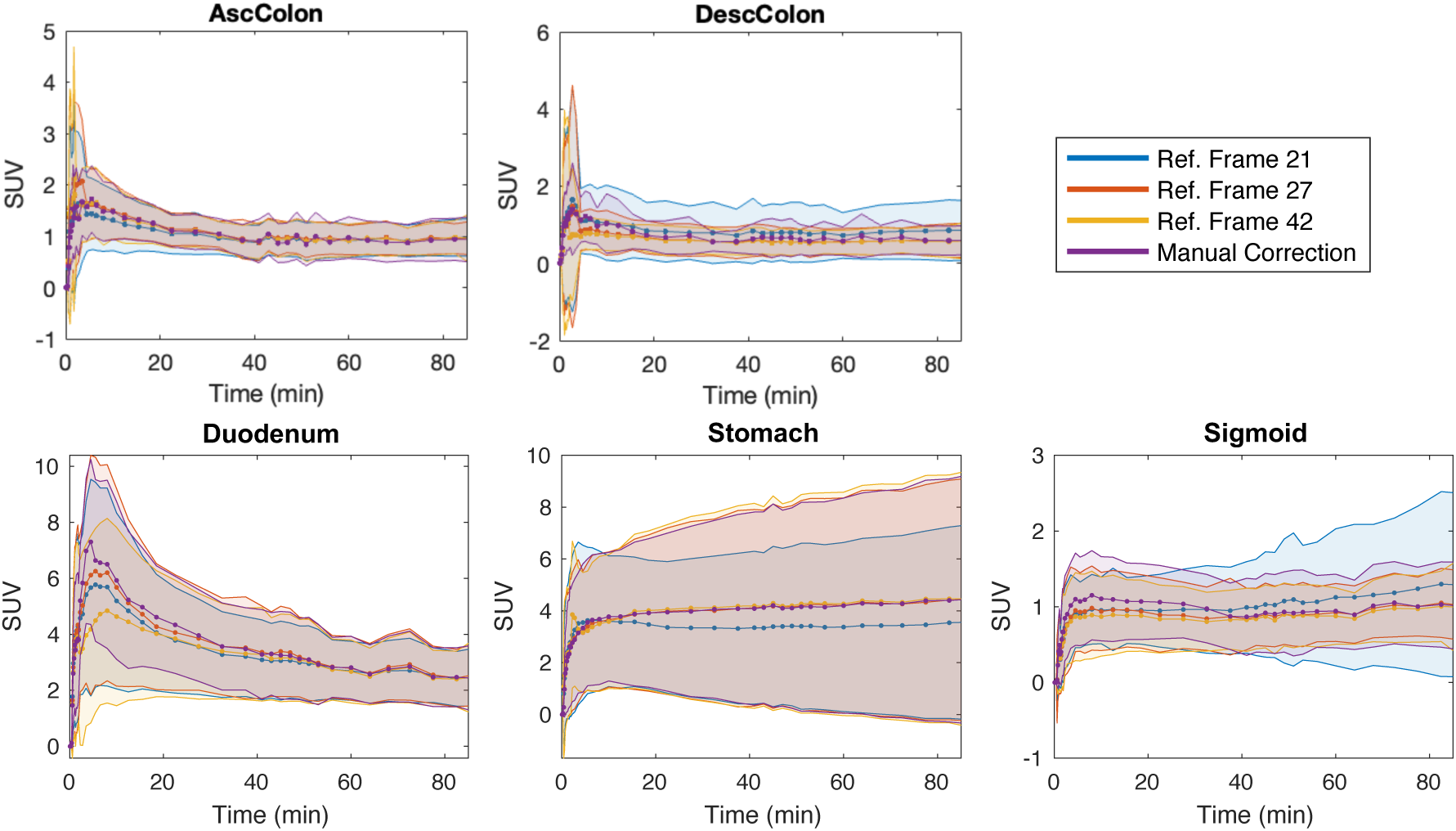
Time activity curves of representative regions across motion correction methods. Data are shown in mean, standard deviation of the VOI. Use of reference frame 42 (in yellow) results in large standard deviations in the ascending colon, descending colon, and duodenum. Frame 21 (11-14 min p.i., in blue) showed deviation from other motion correction reference frames, particularly at the end of the scan in the sigmoid, where in this instance, the bladder overlaps with the sigmoid VOI.

Summed SSR was lowest when frame 27 (35-40 min p.i.) was utilized as the reference frame for semi-automatic motion correction (**Figure 3**). Frame 42, the final frame of the 90-min study, showed the highest summed SSR. As indicated by the heatmaps of organ-level SSR across the different motion corrected reference frames (**Supplemental Figure 1**), myocardium, pancreas, kidneys, and small intestine contributed to disagreement between manual and semi-automatic motion correction methods, with increased SSR when using frame 42 as the reference. These results were generally consistent across conditions.

**Figure 3:**
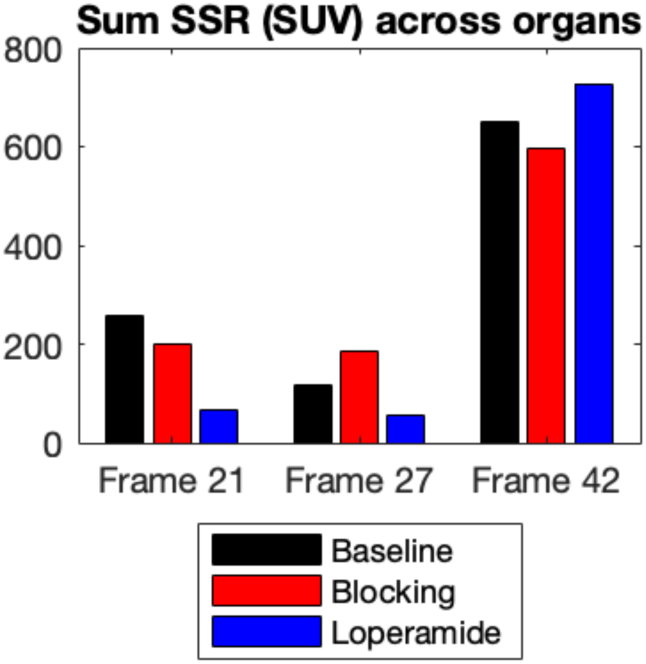
SSR across all organs in a single test subject. Manual motion correction was used as the reference. Each bar represents a single dynamic PET study. Frame 27 (11-14 min p.i.) showed the lowest SSR across all three conditions and was chosen as the reference for further analyses.

With frame 27 as the reference frame, semi-automatic motion correction was performed on the nine additional datasets (3 subjects at baseline, with naloxone and with loperamide). As shown in Table 1, similar SSRs were achieved for two of the three additional subjects across conditions. However, one subject showed a 7-fold higher summed SSR (843.6 vs 116.1) at baseline. For this acquisition, large jumps in the time activity curves were observed in multiple regions (e.g., pancreas, stomach, see **Supplemental Figure 2**) in frames prior to the initiation of motion correction (frame 16, 4-5 min p.i.).

**Table 1:** Summed SSR (units SUV). Each entry is the per-scan SSS. Testing was performed with a single subject with three potential reference frames. Validation was performed with three additional subjects (columns) across the three conditions (rows).

|  |  |  |  |  |  |  |
| --- | --- | --- | --- | --- | --- | --- |
| **Reference frame** | **Test subject** |  |  | **Validation (N = 3)** |  |  |
|  | *Frame 21* | *Frame 27* | *Frame 42* | *Frame 27* | *Frame 27* | *Frame 27* |
| **Baseline** | 258.1 | 116.1 | 651.3 | 843.6 | 20.5 | 69.1 |
| **Naloxone** | 200.4 | 184.5 | 598.0 | 106.3 | 40.9 | 40.1 |
| **Loperamide** | 65.4 | 56.9 | 727.7 | 89.2 | 53.9 | 137.3 |

### Kinetic modeling results with motion correction

Model selection was performed on the final motion-corrected datasets (i.e., both manual and semi-automatic motion-corrected datasets). For all organs except the liver, the 2-tissue, 6-parameter model showed the lowest AIC. The liver was best fit with a 2-tissue, 8-parameter model, which incorporates the hepatic artery and portal vein as dual blood inputs. The slope and Pearson’s correlation coefficients between the kinetic parameters estimated with manual versus frame 27 semi-automatic motion correction are shown in Table 2. Despite the visual TAC mismatches and increased SSR for some organs, overall, the parameter estimates showed a strong relationship (slope range: 0.88-1.02, Pearson’s r > 0.9, p<0.001) for K1, and VTLogan for each of the three conditions. VT, which was derived from compartment modeling, showed still weaker correlations across motion correction methods (slope range: 0.72-1.05, r: 0.52-0.8, p<0.001). Ki also showed the weakest agreement, particularly at baseline (across all conditions: slope range: 0.36-0.43, r: 0.41-0.51, p: 0.001-0.01). This may be due to the liver, which was lower with semi-automatic motion correction than with manual (baseline liver: 0.04 ± 0.01 vs 0.39 ± 0.06, respectively).

**Table 2:** Manual vs semi-automatic motion correction with reference frame 27: Slope, Pearson’s correlation coefficient (r) across individual subjects (N = 4), organs (N = 14)

|  |  |  |  |  |  |  |  |  |  |
| --- | --- | --- | --- | --- | --- | --- | --- | --- | --- |
|  | **Baseline** |  |  | **Naloxone** |  |  | **Loperamide** |  |  |
| **Parameter** | **slope** | **$r$** | **$p$** | **slope** | **$r$** | **$p$** | **slope** | **$r$** | **$p$** |
| $K_1$ | 0.90 | 0.91 | *** | 0.92 | 0.95 | *** | 0.88 | 0.93 | *** |
| $V_T$ | 0.72 | 0.68 | *** | 0.80 | 0.52 | *** | 1.05 | 0.87 | *** |
| $K_i$ | 0.36 | 0.41 | ** | 0.43 | 0.51 | *** | 0.42 | 0.49 | *** |
| $V_{T\text{Logan}}$ | 1.00 | 0.96 | *** | 0.98 | 0.98 | *** | 1.02 | 0.98 | *** |
| TCMDVR | 0.54 | 0.49 | *** | 0.75 | 0.51 | *** | 1.07 | 0.90 | *** |
| LoganDVR | 0.90 | 0.96 | *** | 0.88 | 0.98 | *** | 1.01 | 0.98 | *** |
| ***: $p < 0.001$, **: $p < 0.01$, *: $p < 0.05$ | | | | | | | | | |

### Parameter estimates with manual motion correction

To fully compare motion correction methods, we report parameter estimates for both manual and semi-automatic motion correction. Baseline parameter averages and standard deviations with manual motion correction are reported in Table 3. With the Friedman test, several organ-level parameters showed significant differences across conditions (Table 4). K1 of stomach, sigmoid, and ascending colon were significantly different (p<0.05). With a *post hoc* Tukey test, there was significantly increased K1s with loperamide, relative to naloxone for all three of these regions (p<0.05). Extended tables across all organs and kinetic parameters are included in the supplemental. Logan-based VT (VTLogan) of the descending colon showed differences in means between conditions (p<0.05), but no significant differences when comparing pairwise across conditions. Other parameters showed trends toward significance (e.g., VT of ileum, VTLogan of stomach, and LoganDVR of the ascending colon, kidneys, and ileum). These parameters were generally slightly increased at baseline relative to naloxone and loperamide.

**Table 3:** Average (std. dev.) of kinetic modeling parameter estimates at baseline, manual motion correction (N = 4)

| *Region* | *K1* | *VT* | *Ki* | *VTLogan* | *TCMDVR* | *DVRLogan* |
| --- | --- | --- | --- | --- | --- | --- |
| **AscColon** | 0.105 (0.021) | 3.427 (1.007) | 0.020 (0.003) | 2.300 (1.790) | 1.128 (0.058) | 0.859 (0.682) |
| **DescColon** | 0.082 (0.012) | 5.216 (3.543) | 0.016 (0.002) | 2.012 (0.383) | 1.632 (0.732) | 0.852 (0.308) |
| **Duodenum** | 0.430 (0.072) | 56.816 (56.909) | 0.089 (0.041) | 7.837 (1.462) | 21.359 (22.170) | 3.240 (0.719) |
| **Ileum** | 0.105 (0.043) | 17.121 (11.144) | 0.036 (0.025) | 5.220 (1.823) | 5.339 (2.908) | 2.095 (0.515) |
| **Jejunum** | 0.245 (0.143) | 46.539 (72.601) | 0.084 (0.019) | 6.335 (5.895) | 12.057 (16.427) | 2.781 (2.296) |
| **Kidneys** | 1.322 (0.201) | 6.246 (0.837) | 0.069 (0.014) | 6.294 (0.516) | 2.120 (0.342) | 2.594 (0.342) |
| **Myocardium** | 0.505 (0.198) | 6.886 (1.615) | 0.056 (0.037) | 5.417 (0.933) | 2.373 (0.693) | 2.269 (0.648) |
| **Liver** | 1.036 (0.112) | 25.448 (4.112) | 0.390 (0.060) | 25.209 (2.897) | 7.680 (1.253) | 10.391 (1.582) |
| **Pancreas** | 0.929 (0.110) | 15.014 (8.550) | 0.040 (0.035) | 8.946 (1.551) | 5.336 (3.959) | 3.634 (0.195) |
| **Psoas** | 0.063 (0.063) | 3.038 (0.850) | 0.024 (0.009) | 2.478 (0.525) | - | - |
| **Sigmoid** | 0.049 (0.003) | 15.521 (14.109) | 0.017 (0.001) | 2.682 (0.407) | 4.578 (3.757) | 1.100 (0.161) |
| **Spleen** | 0.908 (0.239) | 5.895 (0.326) | 0.147 (0.149) | 5.800 (0.605) | 2.038 (0.498) | 2.412 (0.509) |
| **Stomach** | 0.232 (0.076) | 21.687 (5.868) | 0.101 (0.041) | 18.388 (7.721) | 7.838 (3.745) | 8.065 (4.383) |
| **TransvColon** | 0.069 (0.029) | 4.514 (2.210) | 0.026 (0.008) | 2.646 (1.014) | 1.519 (0.793) | 1.046 (0.227) |

**Table 4:**
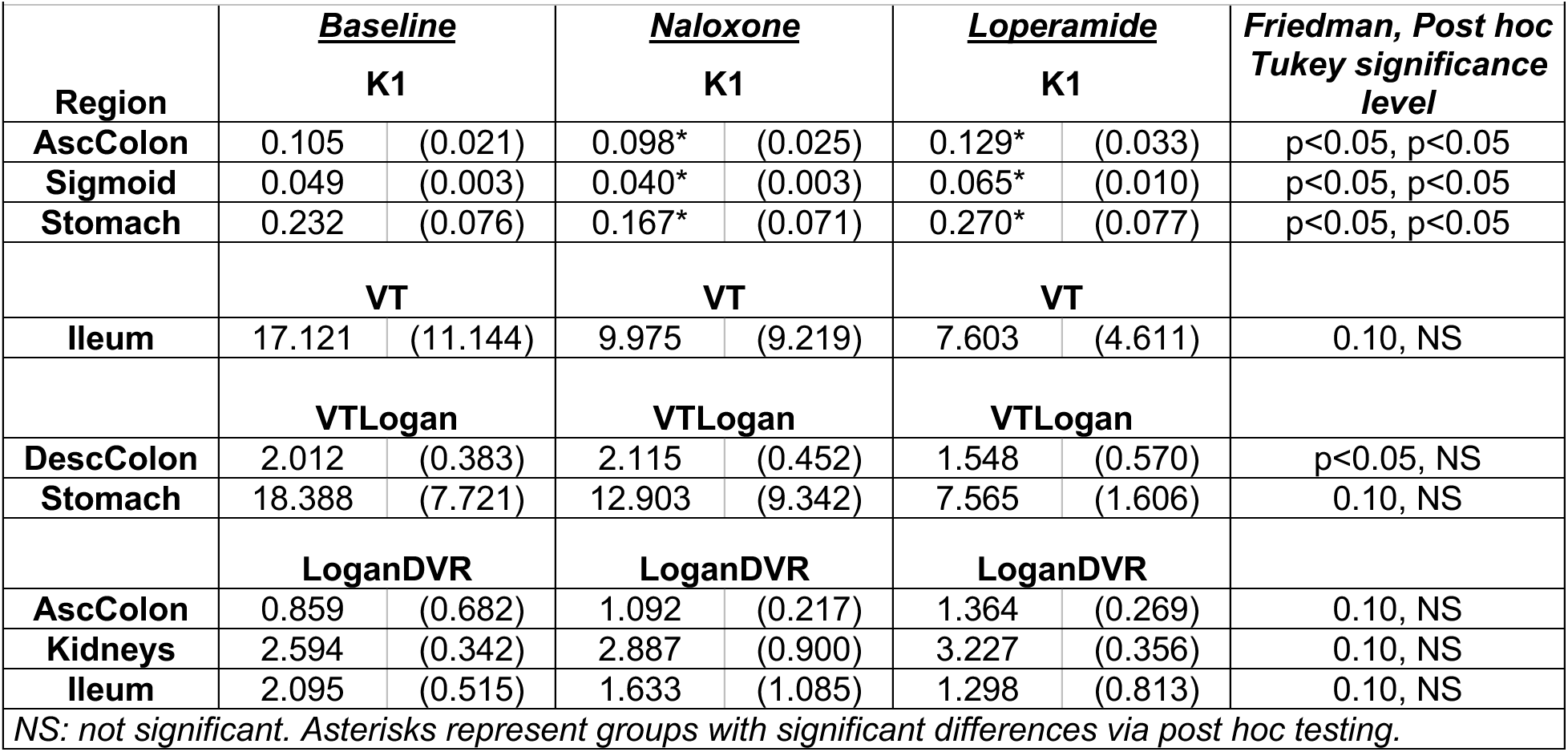
Average (std. dev.) kinetic modeling parameter estimates across conditions with manual motion correction.

### Parameter estimates with semi-automatic motion correction

Despite the high degree of agreement between manual and semi-automatic motion correction in Table 2, we found that there was an overlapping but ultimately distinct set of parameters that were significantly different between conditions, when using semi-automatic motion correction (Table 5). Namely, no K1s were significantly different across the three groups. However, sigmoid VT, ileum VTLogan, descending colon VTLogan, and multiple DVRLogan values (myocardium, duodenum, and ascending colon) all showed statistically significant differences by the Friedman test (p<0.05). Of these, sigmoid VT, ileum VTLogan, and descending colon VTLogan all showed significant decreases with loperamide relative to baseline (p<0.05). However, these parameter estimates also demonstrated a higher coefficient of variation with semi-automatic motion correction, and thus with the small sample size, differences in groups may be governed by outliers.

**Table 5:** Average (std. dev.) kinetic modeling parameter estimates across conditions, semi-automatic motion correction.

|  |  |  |  |  |  |  |  |
| --- | --- | --- | --- | --- | --- | --- | --- |
| Region | <u>Baseline</u><br>K1 |  | <u>Naloxone</u><br>K1 |  | <u>Loperamide</u><br>K1 |  | <u>Significance level</u><br>Friedman, Tukey |
| AscColon | 0.114 | (0.031) | 0.098 | (0.020) | 0.135 | (0.034) | p<0.10, NS |
| TransvColon | 0.072 | (0.031) | 0.103 | (0.049) | 0.192 | (0.211) | p<0.10, NS |
|  | VT |  | VT |  | VT |  |  |
| Sigmoid | 17.755* | (17.303) | 14.645 | (21.556) | 2.882* | (1.042) | p<0.05, p<0.05 |
|  | VTLogan |  | VTLogan |  | VTLogan |  |  |
| DescColon | 3.208* | (0.701) | 2.921 | (0.418) | 1.790* | (0.897) | p<0.05, p<0.05 |
| Ileum | 9.874* | (5.958) | 3.764 | (3.540) | 2.737* | (2.294) | p<0.05, p<0.05 |
| Psoas | 2.713 | (0.621) | 2.560 | (0.686) | 1.876 | (0.092) | 0.10, NS |
| Stomach | 16.834 | (5.991) | 11.480 | (5.941) | 7.955 | (1.796) | 0.10, NS |
|  | LoganDVR |  | LoganDVR |  | LoganDVR |  |  |
| AscColon | 0.939 | (0.474) | 1.166 | (0.179) | 1.622 | (0.385) | p<0.05, NS |
| Duodenum | 2.734 | (1.029) | 4.343 | (1.253) | 3.974 | (0.696) | p<0.05, NS |
| Kidneys | 2.396 | (0.421) | 2.592 | (0.717) | 3.175 | (0.228) | 0.10, NS |
| Myocardium | 2.048 | (0.521) | 1.925 | (0.650) | 2.706 | (0.842) | p<0.05, NS |
| Liver | 9.522 | (1.609) | 10.742 | (3.262) | 12.749 | (0.872) | 0.10, NS |

NS: not significant. Asterisks represent groups with significant differences via post hoc testing.

### Parametric imaging

To assess the performance of motion correction for voxel-wise kinetic modeling, parametric images were generated with and without motion correction for the test subject utilized above. Motion correction was performed with frame 27 as the reference region. As shown in **Figure 4**, visual inspection revealed improved estimation of VTLogan in the GI tract with semi-automatic motion correction (arrows).

**Figure 4:**
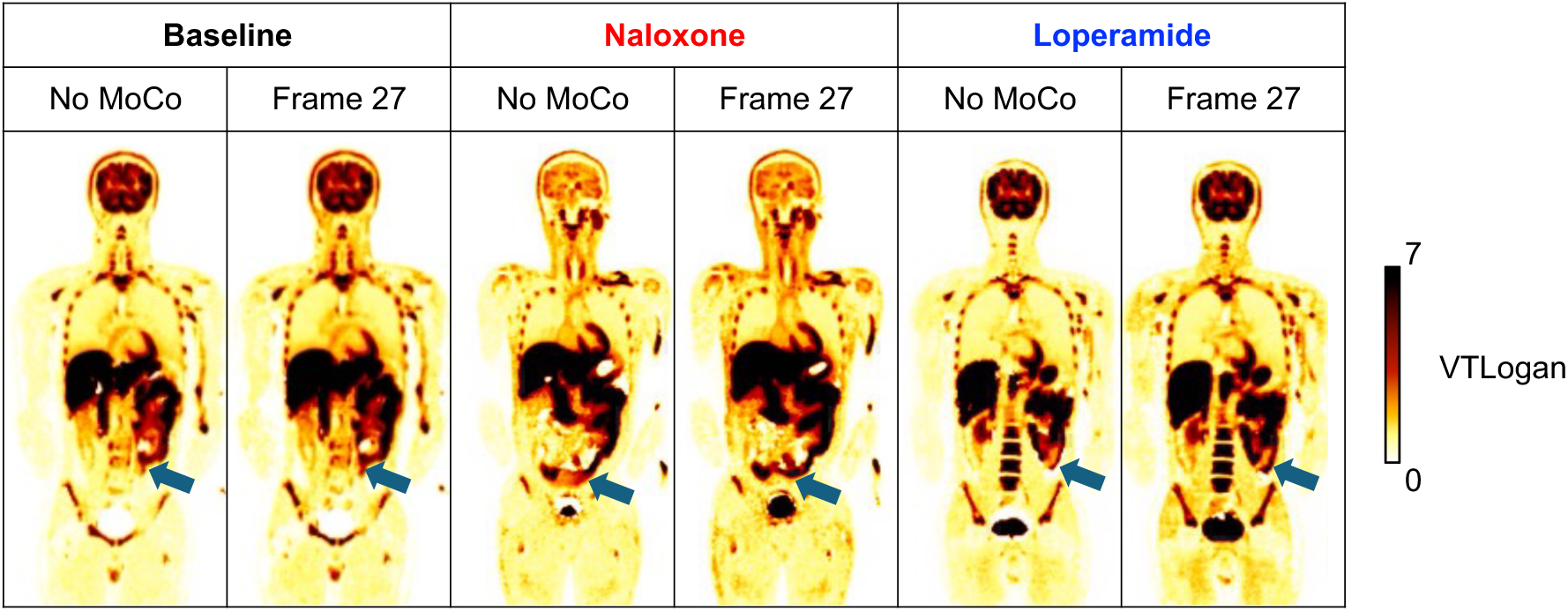
Example parametric images of Logan graphical analysis-based VT (VTLogan). For comparison, VTLogan maps were generated with the original images, without motion correction implemented. Motion-corrected parametric images were generated with frame 27 as the motion correction reference frame. Arrows indicate regions of improved VTLogan estimates with motion correction.

## Discussion

Here, we evaluated [^11^C]-cfn biodistribution in the brain and gastrointestinal system. We found that semi-automatic motion correction was an acceptable strategy for peripheral quantification of [^11^C]-cfn total-body PET. With the framing scheme used in this work, the use of reference frame 27 (35-40 min p.i.) yielded the lowest SSR and a strong correlation (r<0.7) with manual correction. Frame 27 was selected to balance the positioning of the organs due to bladder filling with peak signal in MOR-rich regions in the brain, which was assumed to also be the time of peak SUV in MOR-rich regions in the periphery. While this frame worked well for most studies and improved the parametric VTLogan maps in the GI tract, we note that one baseline acquisition showed high SSR due to motion at timepoints prior to the initiation of motion correction (<4 min p.i.). Because these findings are limited to a small cohort, careful application of these methods in a larger dataset is needed.

As an opioid agonist, loperamide competes with [^11^C]-cfn at peripheral MOR binding sites, while naloxone (a potent antagonist) competes with [^11^C]-cfn both centrally and peripherally. Our results agree with the expected impact of perturbing [^11^C]-cfn binding. The dose level of naloxone delivered in this study is expected to reduce central occupancy by 50% [20], while loperamide was delivered at a standard dose. Loperamide is also expected to act locally within the GI tract, with limited (<2%) circulating bioavailability [21] and thus limited impact on non-GI tract, peripheral organs such as the myocardium. Loperamide is metabolized via CYP3A4, the same cytochrome p450 enzyme as for [^11^C]-cfn [21, 22]. Given that there was no significant impact on plasma curve AUC, we believe that the impact on [^11^C]-cfn metabolism for loperamide studies is limited.

As expected, naloxone administration decreased the thalamic DVR. With manual motion correction, loperamide studies showed greater colon K1, and stomach K1 than naloxone. These results are supported by prior observations that opioids can induce peripheral vasoconstriction, possibly to support increased blood flow to the brain to account for reduced respiration [23], and naloxone has been shown to block opioid receptor-mediated vasodilation in the periphery [24].

While these peripheral differences in K1 were not statistically significant with semi-automatic motion correction, even in retroperitoneal regions (ascending and descending colon) distribution volume measurements (VT, VTLogan, LoganDVR) showed more separation between groups with semi-automatic motion correction, as evident from the greater number of significant Friedman tests when comparing data in Tables 4 and 5. Interestingly, there were no significant differences with *post hoc* testing between baseline and naloxone conditions, but there was separation between baseline and loperamide conditions for sections of the small and large intestines. As expected, some intestinal regions showed statistically significant decreases in distribution volume with loperamide. Specifically, the sigmoid VT, descending colon VTLogan, and ileum VTLogan were significantly decreased relative to baseline with semi-automatic motion correction (**Table 5**).

For graphical modeling in the periphery, we note that the calculation of LoganDVR was accomplished through the quotient of Logan VT’s, rather than direct use of the Logan reference tissue model. This reduces the dependence of DVR on the noise properties of the psoas muscle, which was used as the peripheral reference tissue[8]. Further, it does not require the assumption of a k2’ for each organ across subjects and imaging conditions. Nonetheless for some tissues, with both correction methods there were large coefficients of variation for volume of distribution measurements, particularly for the ileum, stomach, and sigmoid. Based on visual inspection of multiple tissues (including liver, colon regions, and psoas), there were underlying between-subject differences in area under the curve and curve shapes. Such intersubject variability is likely due to physiologic differences in [^11^C]-cfn bioavailability and the presence of tissue radiometabolites across subjects. And while circulating radiometabolites can be accounted for in the blood, no corrections for radiometabolites interacting with tissue were made. If these carbon-11 labeled radiometabolites accumulate in these intestinal regions – which is likely as the GI tract is involved in excretion, the volume of distribution will be inflated. Further, characterization of intestinal wall MOR is challenging, because the wall is only 1-5 mm thick [13] depending on the degree of distension. Whole intestinal sections, as delineated here, will include the intestinal wall, but also include air and solids within the bowel itself. Future work will include the development of *in vivo* correction methods for tissue fraction and tissue-based radiometabolites.

Loperamide acts in the periphery and naloxone acts both centrally and peripherally and are an agonist and antagonist, respectively. Thus, a comparison of loperamide agonism with a peripherally acting antagonist such as methylnaltrexone would be informative. Additionally, the present study is limited to a small number of subjects with a large gap between baseline and naloxone and loperamide acquisitions. However, these results are not only relevant for future [^11^C]-cfn studies but also may be relevant for evaluating other neurotransmitter systems with disrupted brain-GI signaling, such as serotonin, dopamine, and acetylcholine. Disfunction of these neurotransmitter systems has been associated with neuropsychiatric disorders, including depression, Parkinson’s Disease, and long COVID [25–27]. Semi-automatic motion correction of such studies will reduce the methodological burden of image analysis.

## Conclusions

Overall, we found that semi-automatic motion corrections may suffice for quantification in the gastrointestinal tract, though larger sample sizes with additional radiotracers and neurotransmitter systems are needed to confirm these findings. We also found decreased thalamic MOR availability due to naloxone, and decreased volume of distribution of intestinal regions with loperamide, successfully quantifying the MOR brain-GI axis with pharmacologic perturbations of [^11^C]-cfn total-body PET.

## List of abbreviations

Cfn: carfentanil
CNS: central nervous system
CT: computed tompography
GI: gastrointestinal
MOR: mu-opioid receptors
p.i.: post injection
PET: Positron emission tomography
SSR: sum of squared residuals
SUV: standardized uptake value
VOI: volume of interest
VT: total volume of distribution, determined via tissue compartment modeling
VTLogan: total volume of distribution, determined via Logan graphical analysis
DVR: distribution volume ratio
LoganDVR: distribution volume ratio determined via Logan analysis
TCMDVR: distribution volume ratio determined via compartment modeling

## Declarations

### Ethics approval and consent to participate

All procedures performed in studies involving human participants were in accordance with the ethical standards of the institutional and/or national research committee and with the 1964 Helsinki declaration and its later amendments or comparable ethical standards. All individual human participants included in the study have freely given their explicit consent for the processing of their data for one or more specific purposes. All the research studies are performed under IRB (Institutional Review Board) approval within the University of Pennsylvania Human Research Protections Program.

### Consent for publication

Not applicable.

### Data availability

The datasets generated during and/or analyzed during the current study are available from the corresponding author on reasonable request.

### Authors’ contributions

EJ, HK, and JD contributed to the conception and design of the study. Data acquisition was carried out by CH and JC. Analysis was carried out by EL, SL, and JP. The first draft of the manuscript was written by EL, and all authors commented on previous versions of the manuscript. All authors read and approved the final manuscript.

## Acknowledgements

The authors gratefully acknowledge the efforts of the University of Pennsylvania PET Center to acquire these studies.

## References

[1] L. Zhang et al., “Morphine tolerance is attenuated in germfree mice and reversed by probiotics, implicating the role of gut microbiome,” Proceedings of the National Academy of Sciences, vol. 116, no. 27, pp. 13523–13532, 2019, doi: doi:10.1073/pnas.1901182116.

[2] S. Banerjee et al., “Opioid-induced gut microbial disruption and bile dysregulation leads to gut barrier compromise and sustained systemic inflammation,” Mucosal Immunology, vol. 9, no. 6, pp. 1418–1428, 2016/11/01 2016, doi: 10.1038/mi.2016.9.

[3] A. Barkus et al., “The Gut-Brain Axis in Opioid Use Disorder: Exploring the Bidirectional Influence of Opioids and the Gut Microbiome—A Comprehensive Review,” Life, vol. 14, no. 10, p. 1227, 2024. [Online]. Available: https://www.mdpi.com/2075-1729/14/10/1227.

[4] R. S. Hofford, N. L. Mervosh, T. J. Euston, K. R. Meckel, A. T. Orr, and D. D. Kiraly, “Alterations in microbiome composition and metabolic byproducts drive behavioral and transcriptional responses to morphine,” Neuropsychopharmacology, vol. 46, no. 12, pp. 2062–2072, 2021/11/01 2021, doi: 10.1038/s41386-021-01043-0.

[5] E. P. Duffy, R. K. Bachtell, and M. A. Ehringer, “Opioid trail: Tracking contributions to opioid use disorder from host genetics to the gut microbiome,” Neuroscience & Biobehavioral Reviews, vol. 156, p. 105487, 2024/01/01/ 2024, doi: 10.1016/j.neubiorev.2023.105487.

[6] J. G. Dubroff et al., “[(11)C]Carfentanil PET Whole-Body Imaging of mu-Opioid Receptors: A First in-Human Study,” J Nucl Med, May 8 2025, doi: 10.2967/jnumed.124.269413.

[7] E. J. Li et al., “Total-Body Perfusion Imaging with [11C]-Butanol,” J Nucl Med, Aug 31 2023, doi: 10.2967/jnumed.123.265659.

[8] J. G. Dubroff et al., “[11C]Carfentanil PET Whole-Body Imaging of μ-Opioid Receptors: A First in-Human Study,” (in eng), J Nucl Med, May 8 2025, doi: 10.2967/jnumed.124.269413.

[9] F. Awouters, A. Megens, M. Verlinden, J. Schuurkes, C. Niemegeers, and P. A. J. Janssen, “Loperamide,” Digestive Diseases and Sciences, vol. 38, no. 6, pp. 977–995, 1993/06/01 1993, doi: 10.1007/BF01295711.

[10] L. K. S. Sundar et al., “Fully Automated, Semantic Segmentation of Whole-Body 18F-FDG PET/CT Images Based on Data-Centric Artificial Intelligence,“ Journal of Nuclear Medicine, vol. 63, no. 12, pp. 1941–1948, 2022, doi: 10.2967/jnumed.122.264063.

[11] B. Dai et al., “Performance evaluation of the PennPET explorer with expanded axial coverage,” Physics in Medicine and Biology, vol. 68, no. 9, 2023/5// 2023, doi: 10.1088/1361-6560/acc722.

[12] N. Tzourio-Mazoyer et al., “Automated anatomical labeling of activations in SPM using a macroscopic anatomical parcellation of the MNI MRI single-subject brain,” Neuroimage, vol. 15, no. 1, pp. 273–89, Jan 2002, doi: 10.1006/nimg.2001.0978.

[13] M. Macari and E. J. Balthazar, “CT of Bowel Wall Thickening,” American Journal of Roentgenology, vol. 176, no. 5, pp. 1105–1116, 2001, doi: 10.2214/ajr.176.5.1761105.

[14] L. K. Shiyam Sundar et al., “Fully Automated, Fast Motion Correction of Dynamic Whole-Body and Total-Body PET/CT Imaging Studies,” Journal of Nuclear Medicine, vol. 64, no. 7, pp. 1145–1153, 2023, doi: 10.2967/jnumed.122.265362.

[15] J. Logan, J. S. Fowler, N. D. Volkow, G. J. Wang, Y. S. Ding, and D. L. Alexoff, “Distribution volume ratios without blood sampling from graphical analysis of PET data,” J Cereb Blood Flow Metab, vol. 16, no. 5, pp. 834–40, Sep 1996, doi: 10.1097/00004647-199609000-00008.

[16] J. Hirvonen et al., “Measurement of central µ-opioid receptor binding in vivo with PET and [11C] carfentanil: a test–retest study in healthy subjects,” European journal of nuclear medicine and molecular imaging, vol. 36, pp. 275–286, 2009.

[17] J. J. Frost et al., “Multicompartmental analysis of [11C]-carfentanil binding to opiate receptors in humans measured by positron emission tomography,” J Cereb Blood Flow Metab, vol. 9, no. 3, pp. 398–409, Jun 1989, doi: 10.1038/jcbfm.1989.59.

[18] G. Wang, M. T. Corwin, K. A. Olson, R. D. Badawi, and S. Sarkar, “Dynamic PET of human liver inflammation: impact of kinetic modeling with optimization-derived dual-blood input function,” Physics in Medicine and Biology, vol. 63, no. 15, pp. 155004–155004, 2018/7// 2018, doi: 10.1088/1361-6560/aac8cb.

[19] J. Logan et al., “Graphical analysis of reversible radioligand binding from time-activity measurements applied to [N-11C-methyl]-(-)-cocaine PET studies in human subjects,” J Cereb Blood Flow Metab, vol. 10, no. 5, pp. 740–7, Sep 1990, doi: 10.1038/jcbfm.1990.127.

[20] J. K. Melichar, D. J. Nutt, and A. L. Malizia, “Naloxone displacement at opioid receptor sites measured in vivo in the human brain,” European Journal of Pharmacology, vol. 459, no. 2, pp. 217–219, 2003/01/17/ 2003, doi: 10.1016/S0014-2999(02)02872-8.

[21] C. Regnard, R. Twycross, M. Mihalyo, and A. Wilcock, “Loperamide,” J Pain Symptom Manage, vol. 42, no. 2, pp. 319–23, Aug 2011, doi: 10.1016/j.jpainsymman.2011.06.001.

[22] M. G. Feasel, A. Wohlfarth, J. M. Nilles, S. Pang, R. L. Kristovich, and M. A. Huestis, “Metabolism of Carfentanil, an Ultra-Potent Opioid, in Human Liver Microsomes and Human Hepatocytes by High-Resolution Mass Spectrometry,” (in eng), Aaps j, vol. 18, no. 6, pp. 1489–1499, Nov 2016, doi: 10.1208/s12248-016-9963-5.

[23] S. A. Thomas, C. M. Curay, and E. A. Kiyatkin, “Relationships between oxygen changes in the brain and periphery following physiological activation and the actions of heroin and cocaine,” Scientific Reports, vol. 11, no. 1, p. 6355, 2021/03/18 2021, doi: 10.1038/s41598-021-85798-y.

[24] R. A. Cohen and J. D. Coffman, “Naloxone reversal of morphine-induced peripheral vasodilatation,” (in eng), Clin Pharmacol Ther, vol. 28, no. 4, pp. 541–4, Oct 1980, doi: 10.1038/clpt.1980.200.

[25] L. M. T. Dicks, “Gut Bacteria and Neurotransmitters,” (in eng), Microorganisms, vol. 10, no. 9, Sep 14 2022, doi: 10.3390/microorganisms10091838.

[26] A. Mhanna et al., “The correlation between gut microbiota and both neurotransmitters and mental disorders: A narrative review,” (in eng), Medicine (Baltimore), vol. 103, no. 5, p. e37114, Feb 2 2024, doi: 10.1097/md.0000000000037114.

[27] A. C. Wong et al., “Serotonin reduction in post-acute sequelae of viral infection,” Cell, vol. 186, no. 22, pp. 4851–4867.e20, 2023, doi: 10.1016/j.cell.2023.09.013.

