## Supplemental Data for "Brain-gut axis imaging, motion correction with [^11^C]-carfentanil total-body PET"

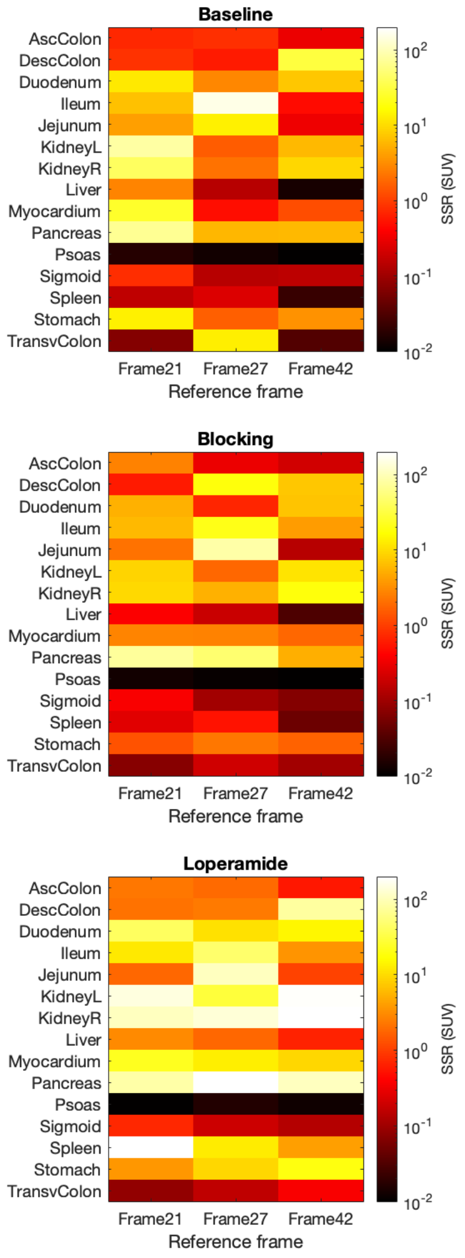

**Supplemental Figure 1:** Regional SSR across motion correction frames. As indicated in Table 1, Frame 27 resulted in the lowest SSR across all three reference frames. Regionally, the pancreas, kidneys, and small intestinal regions contribute to higher SSR for motion correction using frames 21 and 42 as references.

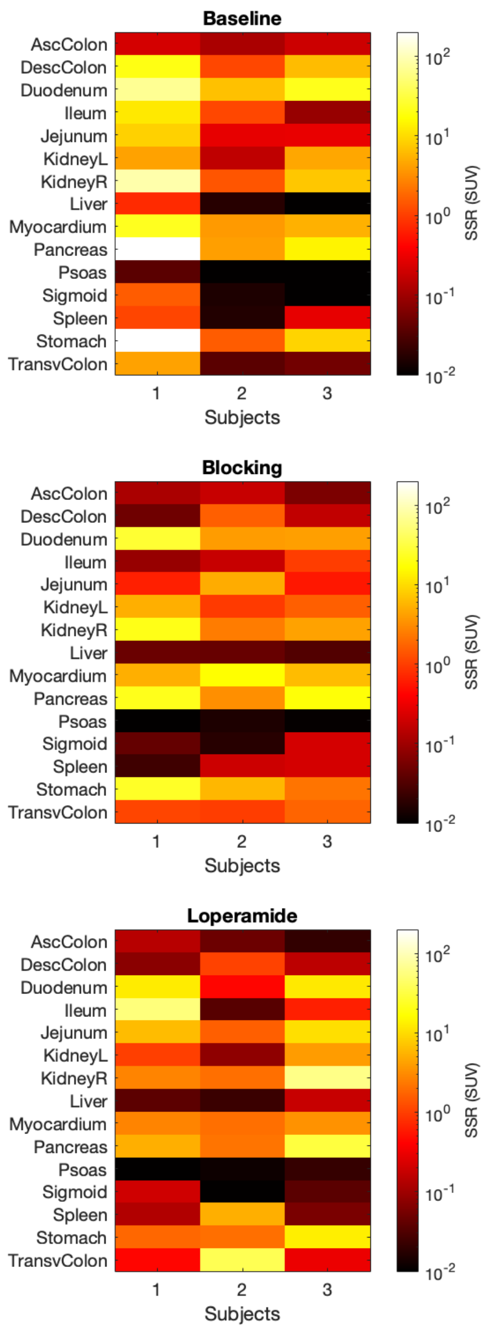

**Supplemental Figure 2:** Regional SSR across additional subjects, conditions when using frame 27 as the reference frame. As indicated in Table 1, Frame 27 resulted in the lowest SSR across all three reference frames. Regionally, the pancreas, stomach, and small intestinal regions contributed to higher SSR.

**Supplemental Data Table 1:** Average (std. dev.) of kinetic modeling parameter estimates at baseline, with naloxone, and with loperamide. Manual motion correction (N = 4).

| Baseline | TCM | K1 | | VT | | Ki | | VTLogan | | TCMDVR | | DVRLogan | |
|---|---|---|---|---|---|---|---|---|---|---|---|---|---|
| Myocardium | 2T6P | 0.505 | (0.198) | 6.886 | (1.615) | 0.056 | (0.037) | 5.417 | (0.933) | 2.373 | (0.693) | 2.269 | (0.648) |
| Liver | 2T8P | 1.036 | (0.112) | 25.448 | (4.112) | 0.390 | (0.060) | 25.209 | (2.897) | 7.680 | (1.253) | 10.391 | (1.582) |
| Stomach | 2T6P | 0.232 | (0.076) | 21.687 | (5.868) | 0.101 | (0.041) | 18.388 | (7.721) | 7.838 | (3.745) | 8.065 | (4.383) |
| Spleen | 2T6P | 0.908 | (0.239) | 5.895 | (0.326) | 0.147 | (0.149) | 5.800 | (0.605) | 2.038 | (0.498) | 2.412 | (0.509) |
| Pancreas | 2T6P | 0.929 | (0.110) | 15.014 | (8.550) | 0.040 | (0.035) | 8.946 | (1.551) | 5.336 | (3.959) | 3.634 | (0.195) |
| Kidneys | 2T6P | 1.322 | (0.201) | 6.246 | (0.837) | 0.069 | (0.014) | 6.294 | (0.516) | 2.120 | (0.342) | 2.594 | (0.342) |
| Duodenum | 2T6P | 0.430 | (0.072) | 56.816 | (56.90) | 0.089 | (0.041) | 7.837 | (1.462) | 21.359 | (22.170) | 3.240 | (0.719) |
| Ileum | 2T6P | 0.105 | (0.043) | 17.121 | (11.14) | 0.036 | (0.025) | 5.220 | (1.823) | 5.339 | (2.908) | 2.095 | (0.515) |
| Jejunum | 2T6P | 0.245 | (0.143) | 46.539 | (72.60) | 0.084 | (0.019) | 6.335 | (5.895) | 12.057 | (16.427) | 2.781 | (2.296) |
| AscColon | 2T6P | 0.105 | (0.021) | 3.427 | (1.007) | 0.020 | (0.003) | 2.300 | (1.790) | 1.128 | (0.058) | 0.859 | (0.682) |
| TransvColon | 2T6P | 0.069 | (0.029) | 4.514 | (2.210) | 0.026 | (0.008) | 2.646 | (1.014) | 1.519 | (0.793) | 1.046 | (0.227) |
| DescColon | 2T6P | 0.082 | (0.012) | 5.216 | (3.543) | 0.016 | (0.002) | 2.012 | (0.383) | 1.632 | (0.732) | 0.852 | (0.308) |
| Sigmoid | 2T6P | 0.049 | (0.003) | 15.521 | (14.10) | 0.017 | (0.001) | 2.682 | (0.407) | 4.578 | (3.757) | 1.100 | (0.161) |
| Psoas | 2T6P | 0.063 | (0.063) | 3.038 | (0.850) | 0.024 | (0.009) | 2.478 | (0.525) | - | - | - | - |
| **Naloxone** | **TCM** | **K1** | | **VT** | | **Ki** | | **VTLogan** | | **TCMDVR** | | **DVRLogan** | |
| Myocardium | 2T6P | 0.539 | (0.196) | 6.293 | (1.486) | 0.021 | (0.010) | 4.858 | (0.926) | 2.115 | (0.609) | 2.216 | (0.718) |
| Liver | 2T8P | 0.991 | (0.054) | 24.172 | (4.446) | 0.463 | (0.085) | 25.916 | (3.418) | 7.390 | (3.140) | 11.920 | (3.653) |
| Stomach | 2T6P | 0.167 | (0.071) | 14.954 | (10.923) | 0.104 | (0.070) | 12.903 | (9.342) | 5.061 | (3.521) | 5.670 | (4.186) |
| Spleen | 2T6P | 0.975 | (0.140) | 5.738 | (0.724) | 0.133 | (0.169) | 5.324 | (0.607) | 2.008 | (0.812) | 2.465 | (0.824) |
| Pancreas | 2T6P | 1.046 | (0.319) | 19.401 | (9.396) | 0.041 | (0.037) | 8.311 | (1.429) | 7.020 | (5.115) | 3.701 | (0.813) |
| Kidneys | 2T6P | 1.437 | (0.393) | 6.353 | (1.074) | 0.066 | (0.017) | 6.284 | (0.924) | 2.166 | (0.661) | 2.887 | (0.900) |
| Duodenum | 2T6P | 0.629 | (0.302) | 23.307 | (21.703) | 0.109 | (0.062) | 11.120 | (1.520) | 7.904 | (7.246) | 5.035 | (1.422) |
| Ileum | 2T6P | 0.125 | (0.063) | 9.975 | (9.219) | 0.034 | (0.030) | 3.885 | (3.077) | 3.364 | (3.031) | 1.633 | (1.085) |
| Jejunum | 2T6P | 0.401 | (0.247) | 12.731 | (3.660) | 0.136 | (0.037) | 12.022 | (2.949) | 4.257 | (1.157) | 5.282 | (1.022) |
| AscColon | 2T6P | 0.098 | (0.025) | 2.961 | (0.807) | 0.021 | (0.003) | 2.481 | (0.595) | 0.977 | (0.217) | 1.092 | (0.217) |
| TransvColon | 2T6P | 0.095 | (0.040) | 3.442 | (1.011) | 0.031 | (0.009) | 3.539 | (1.393) | 1.145 | (0.308) | 1.555 | (0.594) |
| DescColon | 2T6P | 0.071 | (0.025) | 2.673 | (0.267) | 0.024 | (0.008) | 2.115 | (0.452) | 0.926 | (0.319) | 0.985 | (0.433) |
| Sigmoid | 2T6P | 0.040 | (0.003) | 15.171 | (18.235) | 0.016 | (0.003) | 2.867 | (0.844) | 4.014 | (4.021) | 1.274 | (0.399) |
| Psoas | 2T6P | 0.065 | (0.046) | 3.128 | (0.983) | 0.026 | (0.010) | 2.328 | (0.679) | - | - | - | - |
| **Loperamide** | **TCM** | **K1** | | **VT** | | **Ki** | | **VTLogan** | | **TCMDVR** | | **DVRLogan** | |
| Myocardium | 2T6P | 0.533 | (0.249) | 6.395 | (1.707) | 0.062 | (0.029) | 5.108 | (1.352) | 2.812 | (0.667) | 2.786 | (0.926) |
| Liver | 2T8P | 1.108 | (0.124) | 23.180 | (3.163) | 0.410 | (0.105) | 23.837 | (2.666) | 7.480 | (2.473) | 12.811 | (0.754) |
| Stomach | 2T6P | 0.270 | (0.077) | 10.523 | (1.721) | 0.121 | (0.065) | 7.565 | (1.606) | 4.610 | (0.536) | 4.134 | (1.165) |
| Spleen | 2T6P | 1.448 | (0.927) | 5.590 | (0.335) | 0.221 | (0.201) | 5.423 | (0.291) | 2.478 | (0.357) | 2.935 | (0.331) |
| Pancreas | 2T6P | 0.893 | (0.225) | 16.153 | (13.124) | 0.040 | (0.013) | 7.421 | (0.927) | 6.940 | (5.171) | 4.018 | (0.661) |
| Kidneys | 2T6P | 1.522 | (0.286) | 6.139 | (0.764) | 0.074 | (0.036) | 5.960 | (0.228) | 2.708 | (0.353) | 3.227 | (0.356) |
| Duodenum | 2T6P | 0.541 | (0.205) | 35.194 | (49.580) | 0.110 | (0.042) | 6.661 | (2.662) | 14.342 | (19.377) | 3.556 | (1.411) |

| | | | | | | | | | | | | | |
|---|---|---|---|---|---|---|---|---|---|---|---|---|---|
| **Ileum** | 2T6P | 0.101 | (0.017) | 7.603 | (4.611) | 0.026 | (0.020) | 2.506 | (1.678) | 3.229 | (1.669) | 1.298 | (0.813) |
| **Jejunum** | 2T6P | 0.349 | (0.294) | 54.181 | (32.721) | 0.092 | (0.085) | 2.688 | (5.974) | 24.611 | (15.995) | 1.495 | (3.315) |
| **AscColon** | 2T6P | 0.129 | (0.033) | 4.089 | (3.179) | 0.022 | (0.008) | 2.541 | (0.582) | 1.751 | (1.201) | 1.364 | (0.269) |
| **TransvColon** | 2T6P | 0.106 | (0.062) | 3.189 | (0.941) | 0.030 | (0.022) | 2.552 | (0.571) | 1.383 | (0.305) | 1.382 | (0.354) |
| **DescColon** | 2T6P | 0.098 | (0.013) | 3.551 | (1.720) | 0.018 | (0.006) | 1.548 | (0.570) | 1.606 | (0.885) | 0.821 | (0.248) |
| **Sigmoid** | 2T6P | 0.065 | (0.010) | 3.801 | (2.410) | 0.020 | (0.006) | 2.355 | (0.944) | 1.616 | (0.914) | 1.299 | (0.636) |
| **Psoas** | 2T6P | 0.069 | (0.044) | 2.279 | (0.240) | 0.022 | (0.006) | 1.859 | (0.155) | - | - | - | - |

**Supplemental Data Table 2:** Average (std. dev.) of kinetic modeling parameter estimates at baseline, with naloxone, and with loperamide. Semi-automatic motion correction with frame 27 as the reference frame (N = 4).

| **Baseline** | **TCM** | **K1** | | **VT** | | **Ki** | | **VTLogan** | | **TCMDVR** | | **DVRLogan** | |
|---|---|---|---|---|---|---|---|---|---|---|---|---|---|
| **Myocardium** | 2T6P | 0.518 | (0.207) | 7.061 | (1.027) | 0.057 | (0.007) | 5.344 | (0.843) | 2.399 | (0.729) | 2.048 | (0.521) |
| **Liver** | 2T8P | 0.380 | (0.044) | 35.179 | (19.142) | 0.044 | (0.015) | 25.175 | (2.834) | 10.510 | (5.931) | 9.522 | (1.609) |
| **Stomach** | 2T6P | 0.206 | (0.047) | 19.143 | (4.680) | 0.096 | (0.021) | 16.834 | (5.991) | 6.711 | (2.804) | 6.728 | (3.238) |
| **Spleen** | 2T6P | 0.898 | (0.224) | 5.932 | (0.271) | 0.104 | (0.081) | 5.800 | (0.532) | 1.991 | (0.498) | 2.212 | (0.480) |
| **Pancreas** | 2T6P | 1.043 | (0.157) | 8.171 | (1.965) | 0.084 | (0.028) | 8.932 | (1.533) | 2.734 | (0.920) | 3.372 | (0.643) |
| **Kidneys** | 2T6P | 1.374 | (0.198) | 6.264 | (0.852) | 0.065 | (0.008) | 6.318 | (0.593) | 2.067 | (0.400) | 2.396 | (0.421) |
| **Duodenum** | 2T6P | 0.414 | (0.117) | 19.442 | (10.626) | 0.096 | (0.030) | 7.220 | (2.676) | 6.053 | (2.565) | 2.734 | (1.029) |
| **Ileum** | 2T6P | 0.102 | (0.036) | 41.287 | (37.509) | 0.040 | (0.011) | 9.874 | (5.958) | 14.608 | (13.808) | 3.546 | (1.873) |
| **Jejunum** | 2T6P | 0.244 | (0.144) | 49.770 | (71.688) | 0.077 | (0.029) | 6.956 | (6.432) | 12.569 | (14.558) | 2.858 | (2.349) |
| **AscColon** | 2T6P | 0.114 | (0.031) | 6.586 | (6.809) | 0.022 | (0.006) | 2.691 | (1.474) | 2.464 | (2.984) | 0.939 | (0.474) |
| **TransvColon** | 2T6P | 0.072 | (0.031) | 4.432 | (1.863) | 0.028 | (0.008) | 3.470 | (1.668) | 1.384 | (0.287) | 1.248 | (0.386) |
| **DescColon** | 2T6P | 0.171 | (0.068) | 3.502 | (0.996) | 0.018 | (0.003) | 3.208 | (0.701) | 1.173 | (0.455) | 1.203 | (0.236) |
| **Sigmoid** | 2T6P | 0.046 | (0.005) | 17.755 | (17.303) | 0.018 | (0.002) | 1.376 | (1.580) | 5.093 | (4.693) | 0.583 | (0.629) |
| **Psoas** | 2T6P | 0.063 | (0.063) | 3.153 | (0.975) | 0.022 | (0.008) | 2.713 | (0.621) | - | - | - | - |

| **Naloxone** | **TCM** | **K1** | | **VT** | | **Ki** | | **VTLogan** | | **TCMDVR** | | **DVRLogan** | |
|---|---|---|---|---|---|---|---|---|---|---|---|---|---|
| **Myocardium** | 2T6P | 0.520 | (0.164) | 6.484 | (1.193) | 0.047 | (0.030) | 4.641 | (0.964) | 2.292 | (0.962) | 1.925 | (0.650) |
| **Liver** | 2T8P | 0.453 | (0.096) | 22.922 | (3.341) | 0.107 | (0.165) | 25.854 | (3.458) | 6.962 | (2.580) | 10.742 | (3.262) |
| **Stomach** | 2T6P | 0.195 | (0.075) | 29.020 | (34.716) | 0.081 | (0.078) | 11.480 | (5.941) | 9.145 | (9.524) | 4.630 | (2.505) |
| **Spleen** | 2T6P | 0.981 | (0.138) | 5.575 | (0.635) | 0.122 | (0.146) | 5.332 | (0.651) | 1.976 | (0.751) | 2.234 | (0.763) |
| **Pancreas** | 2T6P | 1.061 | (0.356) | 8.839 | (2.200) | 0.064 | (0.034) | 7.998 | (1.411) | 3.182 | (1.605) | 3.216 | (0.654) |
| **Kidneys** | 2T6P | 1.463 | (0.417) | 6.403 | (0.967) | 0.065 | (0.014) | 6.282 | (0.800) | 2.204 | (0.555) | 2.592 | (0.717) |
| **Duodenum** | 2T6P | 0.657 | (0.273) | 11.189 | (1.713) | 0.114 | (0.042) | 10.593 | (1.596) | 3.831 | (0.910) | 4.343 | (1.253) |
| **Ileum** | 2T6P | 0.136 | (0.067) | 13.175 | (8.275) | 0.044 | (0.022) | 3.764 | (3.540) | 4.348 | (2.758) | 1.449 | (1.238) |
| **Jejunum** | 2T6P | 0.326 | (0.142) | 14.988 | (4.554) | 0.112 | (0.016) | 13.458 | (5.080) | 4.927 | (0.677) | 5.236 | (1.078) |
| **AscColon** | 2T6P | 0.098 | (0.020) | 3.452 | (1.311) | 0.022 | (0.004) | 2.958 | (0.755) | 1.128 | (0.216) | 1.166 | (0.179) |
| **TransvColon** | 2T6P | 0.103 | (0.049) | 5.109 | (1.567) | 0.029 | (0.012) | 4.298 | (2.045) | 1.662 | (0.169) | 1.659 | (0.526) |
| **DescColon** | 2T6P | 0.117 | (0.077) | 6.392 | (6.413) | 0.025 | (0.010) | 2.921 | (0.418) | 1.920 | (1.521) | 1.173 | (0.180) |
| **Sigmoid** | 2T6P | 0.042 | (0.009) | 14.645 | (21.556) | 0.018 | (0.002) | 3.313 | (1.484) | 4.135 | (5.493) | 1.331 | (0.615) |

| Psoas | 2T6P | 0.065 | (0.047) | 3.041 | (0.805) | 0.026 | (0.010) | 2.560 | (0.686) | - | - | - | - |
|---|---|---|---|---|---|---|---|---|---|---|---|---|---|
| **Loperamide** | **TCM** | **K1** | | **VT** | | **Ki** | | **VTLogan** | | **TCMDVR** | | **DVRLogan** | |
| **Myocardium** | 2T6P | 0.545 | (0.276) | 6.425 | (1.479) | 0.081 | (0.058) | 5.032 | (1.334) | 2.816 | (0.752) | 2.706 | (0.842) |
| **Liver** | 2T8P | 0.391 | (0.093) | 21.583 | (2.386) | 0.111 | (0.150) | 23.974 | (2.722) | 6.898 | (1.310) | 12.749 | (0.872) |
| **Stomach** | 2T6P | 0.265 | (0.075) | 11.002 | (2.566) | 0.118 | (0.068) | 7.955 | (1.796) | 4.742 | (1.107) | 4.270 | (1.096) |
| **Spleen** | 2T6P | 1.365 | (0.784) | 5.795 | (0.320) | 0.125 | (0.094) | 5.401 | (0.307) | 2.541 | (0.482) | 2.888 | (0.279) |
| **Pancreas** | 2T6P | 0.911 | (0.222) | 15.013 | (14.381) | 0.059 | (0.021) | 7.140 | (1.187) | 6.032 | (4.884) | 3.821 | (0.729) |
| **Kidneys** | 2T6P | 1.538 | (0.298) | 6.015 | (0.987) | 0.068 | (0.022) | 5.940 | (0.168) | 2.593 | (0.296) | 3.175 | (0.228) |
| **Duodenum** | 2T6P | 0.548 | (0.197) | 35.519 | (48.969) | 0.106 | (0.036) | 7.457 | (1.363) | 14.627 | (19.769) | 3.974 | (0.696) |
| **Ileum** | 2T6P | 0.111 | (0.020) | 24.768 | (41.234) | 0.041 | (0.009) | 2.737 | (2.294) | 9.362 | (14.807) | 1.431 | (1.165) |
| **Jejunum** | 2T6P | 0.343 | (0.299) | 61.579 | (33.317) | 0.087 | (0.071) | 1.652 | (8.378) | 28.217 | (17.184) | 0.897 | (4.434) |
| **AscColon** | 2T6P | 0.135 | (0.034) | 2.804 | (0.369) | 0.024 | (0.009) | 3.028 | (0.647) | 1.232 | (0.285) | 1.622 | (0.385) |
| **TransvColon** | 2T6P | 0.192 | (0.211) | 9.988 | (13.958) | 0.021 | (0.006) | 3.680 | (2.026) | 4.615 | (6.729) | 1.945 | (1.008) |
| **DescColon** | 2T6P | 0.162 | (0.080) | 2.345 | (0.490) | 0.021 | (0.006) | 1.790 | (0.897) | 1.030 | (0.310) | 0.961 | (0.501) |
| **Sigmoid** | 2T6P | 0.062 | (0.010) | 2.882 | (1.042) | 0.021 | (0.005) | 2.379 | (1.001) | 1.224 | (0.353) | 1.289 | (0.619) |
| **Psoas** | 2T6P | 0.069 | (0.045) | 2.329 | (0.339) | 0.022 | (0.007) | 1.876 | (0.092) | - | - | - | - |
